# A model to forecast regional demand for COVID-19 related hospital beds

**DOI:** 10.1101/2020.03.26.20044842

**Authors:** Johannes O. Ferstad, Angela Gu, Raymond Y. Lee, Isha Thapa, Andrew Y. Shin, Joshua A. Salomon, Peter Glynn, Nigam H. Shah, Arnold Milstein, Kevin Schulman, David Scheinker

## Abstract

COVID-19 threatens to overwhelm hospital facilities throughout the United States. We created an interactive, quantitative model that forecasts demand for COVID-19 related hospitalization based on county-level population characteristics, data from the literature on COVID-19, and data from online repositories. Using this information as well as user inputs, the model estimates a time series of demand for intensive care beds and acute care beds as well as the availability of those beds. The online model is designed to be intuitive and interactive so that local leaders with limited technical or epidemiological expertise may make decisions based on a variety of scenarios. This complements high-level models designed for public consumption and technically sophisticated models designed for use by epidemiologists. The model is actively being used by several academic medical centers and policy makers, and we believe that broader access will continue to aid community and hospital leaders in their response to COVID-19.

**LINK TO ONLINE MODEL:** https://surf.stanford.edu/covid-19-tools/covid-19/

## INTRODUCTION

The US classified the coronavirus disease pandemic (COVID-19) as a national emergency on March 14, 2020. Cumulative US cases surged beyond 122,000 on March 29, 2020. [2, 3] Its rapid spread in China and Italy quickly overwhelmed available hospital beds. [4, 5] County-level forecasts of demand for hospital beds based on data commonly available to hospitals would help guide US hospital efforts to anticipate and mitigate similar bed shortages. [6, 7]

In order to plan their response, hospital and public health officials need to understand how many people in their area are likely to require hospitalization for COVID-19; how these numbers compare to the number of available intensive care and acute care beds; and how to project the impact of social-distancing measures on utilization. Since the majority of people with COVID-19 are asymptomatic and the rates of cases requiring hospitalization differ significantly across age groups, answering these questions requires understanding the epidemic and accounting for the specific vulnerabilities of the local population. [8, 9] The numbers of people in each age group differ significantly across US counties, as do the available hospital resources [10, 11]. Initial analyses of the potential impact of COVID-19 have spurred governmental action at the federal and state level; however, significant differences remain in the policies implemented across states and counties. [9, 12]

In order to help facilitate adequate and appropriate local responses, we developed a simple model to project the number of people in each county in the United States who are likely to require hospitalization as a result of COVID-19 given the age distribution of the county per the US Census. The model compares the projected number of individuals needing hospitalization to the publicly known numbers of available intensive and acute care beds and allows users to model the impact of social-distancing or other measures to slow the spread of the virus. The uncertainty surrounding the numbers of people infected and the rates of spread make it difficult to evaluate the accuracy of projections generated by complex epidemiological models. The model presented errs on the side of simplicity and transparency to allow non-specialist policymakers to fully understand the logic and uncertainty associated with the estimates.

## THE PROJECTION MODEL

For each US county, the model accepts as an input the number of COVID-19 hospitalizations and the associated doubling time, if these are available. If these are not available, the model imports the latest number of confirmed cases from the New York Times online repository and accepts user-entered parameters of the ratio of total cases to confirmed cases (e.g., 5:1) [8, 23] and the COVID-19 population-level doubling time (e.g., 7 days) [13]. The effects of interventions that mitigate the spread of infection (such as social distancing) are simulated with user-entered parameters in the form of a greater doubling time and a start date for that new doubling time. County-specific hospitalization rates are derived from combining age-distributions derived from the US census [11] and age-group specific estimates of the case rates of severe symptoms, critical symptoms, and mortality (together morbidity) derived from Imperial College COVID-19 Response Team [9]. The default assumptions are that: people are admitted to the hospital on the day they test positive (the assumptions will change when testing begins for non-symptomatic people); those with severe and critical symptoms spend, respectively, 12 days in acute care and 7 days in intensive care; and 50% of each type of bed is available for COVID-19+ patients. [14, 15] The numbers of patients requiring each type of bed are compared to the numbers of relevant beds derived from data from the American Hospital Association. [10] The detailed technical description of the model is available in our Technical Supplement.

To facilitate use by hospital and public health officials, the model is deployed through an interactive online website that allows users to generate dynamic, static, and spatial estimates of the number and rate of severe, critical, and mortality case rates for each county or group of counties. These data are displayed along with the number of intensive care and acute care hospital beds in the corresponding region.

The urgency of the challenge and widespread data sharing have led to the development of numerous models of COVID-19: models designed to share the most recent data on confirmed cases and deaths [16]; technical models intended for use by epidemiologists [17, 18, 21]; and models presenting high-level summaries of the potential for COVID-19 associated bed demand to overwhelm hospital capacity [9, 19, 22]. The model presented here fits the needs of local hospital and government leaders, many of whom lack access to trained epidemiologists or data analysts. The website allows such leaders to study projections of the spread of COVID-19 based on their county-level hospitalization information, if these are available, and otherwise presents projections based on data from the rest of the US and assumptions from the literature. The model, deployed as a website, is relatively easy to understand as the underlying dynamics are specified in terms of assumptions interpretable to those with no specialist training.

As stakeholders gather data about the spread of COVID-19 in their region, the model permits them to tailor inputs and generate new estimates in real time. As new epidemiological data become available, the model is updated by those maintaining it with new age-specific rates of severe disease, critical disease, mortality and their associated LOSs. For example, the first update was from case-rates derived from data on Wuhan [20] to the case-rates derived from the Imperial report.

As expected, the model predicts US counties with large, older populations are likely to have the highest demand for hospital beds. Counties with a high number of confirmed cases are likely to be first to see the demand for beds exceed hospital bed availability. Furthermore, smaller counties with relatively few hospital beds are likely to see the demand for beds exceed hospital bed availability at lower fractions of the population being COVID-19+ (see Table 1). Local leaders in these vulnerable communities should pay particular attention to the case growth rate if they seek to tailor their response rapidly to predicted downstream resource constraints.

**TABLE 1:** County Utilization with 1% of Population Symptomatic.

| **cover** | **County** | **Population** | **Reported Cases on March 26** | **Number of Acute Beds** | **Number of ICU Beds** | **Acute Bed Utilization if 1% Symptomatic** | **ICU Bed Utilization if 1% Symptomatic** |
| --- | --- | --- | --- | --- | --- | --- | --- |
| California | Santa Cruz County | 273,765 | 25 | 68 | 6 | 199% | 936% |
| Maryland | Harford County | 251,025 | 5 | 53 | 6 | 245% | 917% |
| Michigan | Clinton County | 77,896 | 6 | 23 | 2 | 176% | 868% |
| Massachusetts | Plymouth County | 512,135 | 101 | 131 | 16 | 211% | 763% |
| Tennessee | Hawkins County | 56,402 | 1 | 16 | 2 | 202% | 734% |
| New Mexico | Chaves County | 65,459 | 3 | 18 | 2 | 172% | 722% |
| New Hampshire | Rockingham County | 305,129 | 56 | 64 | 10 | 265% | 708% |
| Minnesota | Dakota County | 418,201 | 21 | 109 | 12 | 185% | 660% |
| Massachusetts | Hampshire County | 161,159 | 11 | 34 | 6 | 233% | 590% |
| Ohio | Delaware County | 197,008 | 12 | 43 | 6 | 216% | 590% |
| Wisconsin | St. Croix County | 87,917 | 4 | 45 | 3 | 95% | 553% |
| Alabama | Baldwin County | 208,107 | 4 | 95 | 10 | 122% | 544% |
| North Carolina | Chatham County | 69,791 | 6 | 21 | 4 | 200% | 539% |
| Arizona | Santa Cruz County | 46,584 | 1 | 16 | 2 | 142% | 539% |
| Florida | Monroe County | 76,325 | 11 | 21 | 4 | 225% | 538% |
| Massachusetts | Norfolk County | 698,249 | 222 | 328 | 31 | 110% | 534% |
| North Carolina | Rowan County | 139,605 | 6 | 162 | 6 | 45% | 529% |
| Georgia | Coweta County | 140,516 | 10 | 37 | 5 | 183% | 512% |
| Iowa | Kossuth County | 15,075 | 1 | 18 | 1 | 48% | 495% |
| California | Placer County | 380,077 | 30 | 146 | 20 | 140% | 489% |

Our results suggest, in line with basic epidemiological principles, that hospitalization rates are very sensitive to contagion doubling time. Despite the uncertainty about doubling time, the model demonstrates that across a large set of assumptions, efforts to increase hospital bed capacity are not on their own likely to be sufficient to ensure that hospital capacity meets demand.

When considering the limited healthcare resources anticipated, the need for strengthening collaborative operational planning and research between institutions and across the public and private sectors is paramount. While hospital leaders may be concerned with the impact of COVID-19 infections at their facility, our model assesses the potential impact of the epidemic at a county and state level. Assessing the regional impact can permit collaborative and coordinated planning between public health and hospital leaders facing an unprecedented challenge. Regional and state leaders are better equipped to understand the potential implications, such as funding and cost-sharing, of this epidemic across counties within a state. These conversations are facilitated by the ability to study several counties in a region.

One of the greatest challenges to continuously adjusting regional policies best tailored to meet the challenge of COVID-19 is that healthcare and political leaders lack familiarity with the concept of exponential growth of a highly transmissible deadly pathogen—particularly one with a 3-to 14-day lag between infection and needing a hospital bed. Forecasting demand for hospital resources a week to a few weeks ahead helps address that handicap.

There are several limitations to the model. First, projections related to the spread of a pandemic at this early stage are plagued with uncertainty about critical model parameters, e.g., the doubling time and the true number of cases in the population. To account for this, the model was designed to produce projections given current assumptions about parameters (with the option to update those variables as better data become available). Second, although estimates of disease propagation follow first-order evidence, important input parameters may not have been considered in the model which may affect hospitalization rates and bed availability. For example, assumptions on shared healthcare resources (such as the use of pediatric beds for adult hospitalization overflow) are not modifiable by users but only by those maintaining the model. Finally, the model estimates the effect of one or the summation of more than one intervention (by modification of the doubling time) but does not ascribe the individual impact of sequential interventions.

In this report, we describe an online, real-time, interactive simulation model to facilitate local policy making and regional coordination by providing estimates of hospital bed demand and the impact of measures to slow the spread of the infection.

## Data Availability

The data used in this work is publicly available.

https://surf.stanford.edu/covid-19-tools/covid-19/

## ACKNOWLEDGEMENTS

We thank Grace Lee, Teng Zhang, Jacqueline Jil Vallon, and Francesca Briganti for their help with this work. We also thank Amber Levine for her help testing and improving the tool.

---

**FIGURE 1:**
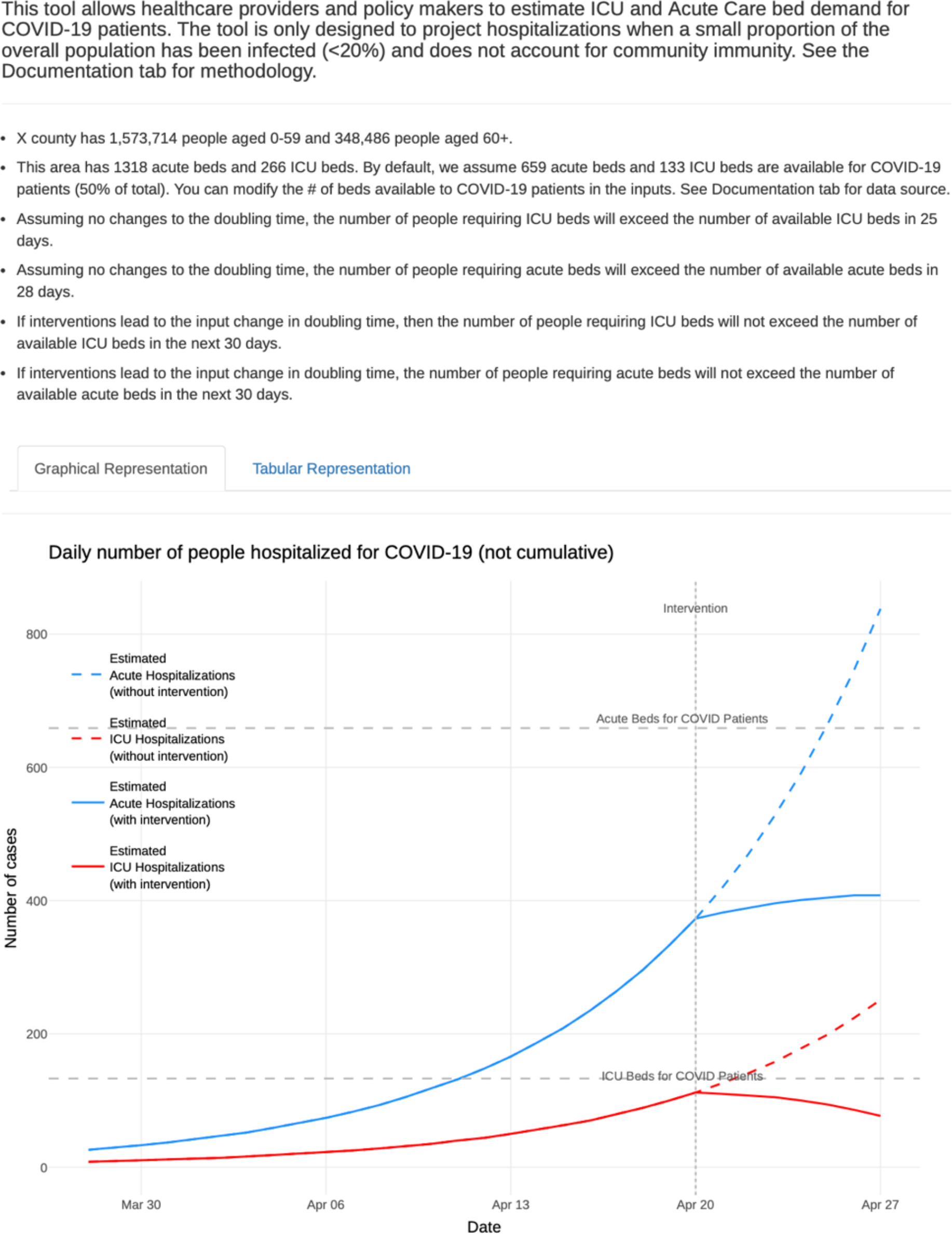
Example output showing the model output simulating an intervention that changes the growth rate of COVID-19 hospitalizations on April 20. The plot is a hypothetical situation and not a forecast for any county.

**FIGURE 2:**
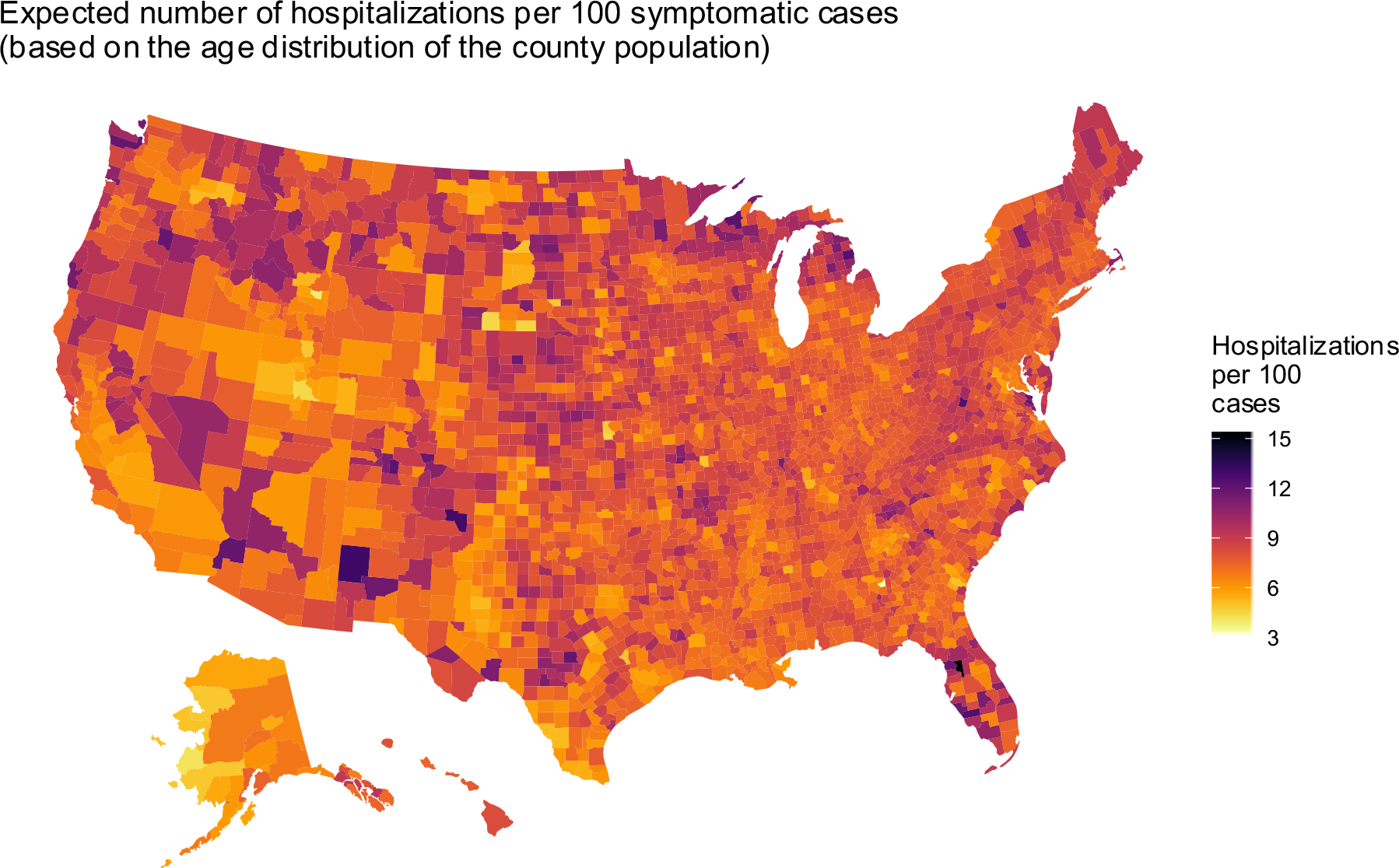
US Map

## Technical Supplement

### Calculation of the Number of COVID-19+ People

The model accepts as an input the number of COVID-19 hospitalizations and the associated doubling time, if these are available. If these are not available, for each US county, the model imports the latest total number of confirmed cases from the New York Times online repository and accepts user-entered parameters of the ratio of total cases to confirmed cases (e.g., 10:1) [1, 9] and the COVID-19 population-level doubling time (e.g., 7 days) [2]. Following a simple exponential model, the total number of COVID-19+ people on each subsequent day N is the product of the initial number of total cases and 2 to the power given by N divided by the doubling time input parameter. Users can simulate the effects of interventions that mitigate the spread of infection (such as social distancing) by entering new doubling times and dates for these interventions to take effect. If these are input, the number of COVID-19+ people on subsequent days is calculated using the new doubling time with the previous formula. Users are referred to online tools to estimate changes in doubling time associated with the impact of social-distancing interventions. [3]

### Calculation of Hospitalization Rates

County-specific age distributions are derived from the US census [4] and age-group specific estimates of the case rates of severe symptoms, critical symptoms, and mortality derived from Imperial College COVID-19 Response Team [5]. For each county, the proportion of each age group relative to the total population is calculated. Severe and critical symptom case-rates for each age group are weighted by these proportions, and the hospitalization rate is calculated as the sum of these weighted severe and critical symptom case-rates.

### Calculation of Hospitalizations and Hospital Capacity

The model includes parameters for the hospital length of stay for COVID-19+ patients with severe and critical symptoms set to, respectively, 12 days in acute care and 7 days in intensive care. By default, 50% of the available acute and ICU beds is shown as the overall capacity for COVID-19+ patients. But another parameter allows the user to adjust the number of each type of bed that is available for COVID-19+ patient. [6, 7] The number of patients with severe and critical symptoms requiring hospitalization each day is calculated assuming that patients are admitted to the hospital on the day they test positive (this assumption will change when testing begins more seriously for non-symptomatic people) and are discharged after their length of stay. The cumulative number of patients with severe and critical symptoms requiring hospitalization is estimated as the product of the corresponding hospitalization rate and cumulative number of COVID+ people in the population. The number of patients in hospital beds on a specific day is defined as the cumulative number of patients admitted up to that day minus the corresponding cumulative number of patients discharged.

Hospital capacity at county level is based on the number of hospital beds in each US county from the American Hospital Association. The acute care beds include general medical, surgical, and long-term acute care beds for adults. The intensive care unit beds include all adult intensive care unit beds other than neonatal [8]. As discussed above, the number of hospital beds available for COVID-19+ patients is by default 50% of overall beds, but can be adjusted by the user.

